# Research on the Influence of Effective Distance Between Cities on the Cross-regional Transmission of COVID-19

**DOI:** 10.1101/2020.03.27.20044958

**Authors:** Shanlang Lin, Yanning Qiao, Junpei Huang, Na Yan

**Author notes:** Correspondence to: School of Economics and Management, Tongji University, China.

## Abstract

The COVID-19 epidemic in China has been effectively controlled. It is of great significance to study the law of cross-regional spread of the epidemic, for the prevention and control of the COVID-19 in the future in China and other countries or regions. In this study, the cross-regional connection intensity between cities was characterized based on the probability and the effective distance of the shortest path tree, and the empirical analysis was carried out based on the high-frequency data such as the cases of COVID 19 outbreaks. It is concluded that the higher the intensity of inter-city connection, the larger scale the cross-regional spread of the epidemic.

## 1 Introduction

In December 2019, the novel coronavirus (COVID-19) occurred in Wuhan and quickly spread to other provinces. According to statistics, as of March 25, 2020, a total of 81,285 confirmed cases, 3,287 deaths, and 3,947 confirmed cases have been reported in mainland China. Among them, a total of 67,801 confirmed cases in Hubei Province (50006 cases in Wuhan), 3169 death cases (2531 cases in Wuhan), and 3431 confirmed cases (3407 cases in Wuhan). The cumulative number of confirmed cases in Hubei Province accounted for 83.7% of mainland China, and the number of deaths accounted for 96.4% of Mainland China, of which Wuhan accounted for 61.7% and 76.9% of Mainland China respectively. The epidemic has been contained in China, and there are basically no new cases in the country, or only cases associated with imported cases abroad in some regions. The focus has been shifted to preventing imported cases. However, the COVID-19 is still serious in countries outside China. According to the data reported by the WHO, as of 10:00 Central European Time on the 25th, there were 332,331 confirmed cases in countries outside China, an increase of 40,611 cases were reported that day; a total of 15,153 death cases, an increase of 2,198 cases that day. Among them, 69,176 cases were confirmed in Italy, 51,914 in the United States, 39,673 in Spain, 31,554 in Germany, and 24,811 in Iran. Therefore, studying prevention and control measures against the COVID-19 and the effects of it in China has certain reference and inspiration value for countries outside China.

Looking back at the outbreak of the COVID-19 epidemic and its prevention and control, Wuhan was the first to report 27 confirmed cases on December 31, 2019, and to China on January 22, 554 confirmed cases, including 444 in Hubei Province. The week starting on January 24 is the traditional Chinese spring festival holiday, which is the period with the largest turnover of people. In 2019, more than 421 million people were transported by commercial transportation agencies, including trains, cars, and aviation. This brings difficulties to COVID-19 prevention and control. On January 23, Wuhan announced the closure of the city, and the city’s urban bus, subway, ferry, and long-distance passenger transportation were suspended; for no special reason, citizens should not leave Wuhan, and the leaving-Wuhan corridors in airport and railway station are temporarily closed. On January 23, Zhejiang, Guangdong, and Hunan provinces initiated a level I response. On January 25, all provinces except Qinghai and Tibet initiated a level I response. On January 30, Tibet province launched a level I response as the last province. This is the highest level of emergency response adopted in accordance with China’s National Emergency Plan for Public Health Emergencies, which classifies public health emergencies into particularly major (level I), major (level II), large (level III) and general (level IV) level 4. According to the plan of level I response, the provincial government is responsible for emergency response, adopts preventive control measures in time, adopts emergency mobilization and collection of relevant personnel, materials, vehicles, related facilities, and equipment, adopts isolated sites and blocks epidemic areas to ensure various supplies needed in emergency response. But before the “lockdown” of Wuhan, according to statistics, millions of citizens have left Wuhan, and they may be the main source of the COVID-19 infection in other parts of China. Therefore, it is of a certain value to analyze the impact of Wuhan’s intercity connection with other regions on the spread of the epidemic and the role of local prevention and control measures.

Observe the distribution of the COVID-19 confirmed cases in various regions of Hubei Province and other provinces (cities and districts) in China. The number of cases did not increase with the proximity of Wuhan, even in Hubei province. Then, an important question is that the cities have different degrees of economic and personnel links with Wuhan. Does the link have an internal relationship with the spread of the epidemic? The public health policies for COVID-19 prevention and control in different regions are not exactly the same, and the degree of strictness also varies. It is also an important question of how such prevention and control measures are matched with the degree of spread.

This study has several strengths. First, based on the theory of infectious diseases, the theory of complex network system and the characteristics of COVID-19, considering that the incubation period was still infectious, we propose an epidemic model which describes how people move between cities and spread disease; Second, the concept of “effective distance”, which is not related to geographical distance, but related to transportation and population size, is introduced to analyze the effect of effective distance on epidemics and the transmission mechanism of the COVID-19; Third, big data and data mining technologies are used to retrieve the database of population migration data, Baidu Index and so on. Fourth, by applying the methods of descriptive statistics, multivariate analysis, econometrics, and the mathematical modeling methodology, we verified the effect of prevention and control efforts on reducing the transmission of COVID-19 among cities.

The rest of the paper is organized as follows. Section 2 presents the literature review of the mathematical model of the epidemic transmission and how travel can contribute to the rapid spread of disease; Section 3 introduces a mathematical model of how COVID-9 flows and spreads disease between different cities; Research and assumption on model constructing and the meaning of each variable in the study and their sources are introduced in Section 4. Section 5 shows the empirical analysis result and combs the transmission mechanism of COVID-19; In the last section, we make a conclusion and discussion of the content and proposes the direction of future research.

## 2 Literature Review and Hypothesis

### 2.1 Literature Review

The epidemic of infectious diseases is a complex spreading process that occurs in population. There is a long history of modeling infectious disease epidemics (Anderson et al., 1992), and various modeling paradigms have been developed. Kermark & Mckendrick (1927) proposed the classic SIR model, which is a compartmental model. Assuming that every individual is the same, the population is homogeneous mixing. The contact is instant and independent of history, infection rate and recovery rate are constant. All people in the same state form a compartment, and as the state changes, personnel move between the compartments. With the growth of urban population and the development of transportation networks, social mobility has increased, and the spatial expansion of infectious diseases has shown a new pattern. Especially when people move between different regions, the spread of infectious diseases is very common. Understanding the impact of human movement patterns on the prevalence of infectious diseases has attracted considerable attention (Gonzalez et al., 2008), and a meta-population model derived from ecology has been applied in the field of infectious diseases. With the rise of complexity science, the micro-modeling specification has been developed and combined with social networks, a network-based micro-individual modeling method has been developed, which provides a new way to understand the spread of infectious diseases. These models enable epidemiologists and health authorities to understand the transmission process, predict its impact on healthy populations, and assess the effectiveness of different mitigation and prevention strategies. Since 2002, the coronaviruses (CoVs) have emerged as major global health threats, namely severe acute respiratory syndrome coronavirus (SARS-CoV; in 2002) that spread to 37 countries, and Middle East respiratory syndrome coronavirus (MERS-CoV; in 2012) that spread to 27 countries.

Part of the literature studied the treatment of emerging infectious diseases by medical staff to reduce or avoid transmissions, such as Kisting et al. (2012), Chea et al. (2014), Barbisch et al. (2015), Koening (2015) and others. Another part of the literature studied public health policy issues for infectious diseases. Wilder-Smith (2020) and Huremović (2019) analyzed and compared the clear definitions and scope of common public health policies, including isolation, quarantine, social distancing and so on, and other literature such as Rothstein & Talbott (2007), Wang et al. (2012), Sakaguchi et al. (2012). After the outbreak of COVID-19 in Wuhan, most of the research focused on estimating the source, transmission characteristics and the estimation of the scale (Mizumoto et al., 2020; Zhou et al., 2020; Hébert-Dufresne et al., 2020; Liu et al., 2020; Fu et al., 2020; Yeo et al., 2020). In particular, the basic reproduction ratio has been widely debated as a parameter that reflects the speed of virus transmission. Some literatures analyzed the spread of COVID-19 using population migration data, which are calculated based on the migration outflow from Wuhan (Zhan et al., 2020; Ding et al., 2020; Zheng et al., 2020; Chen et al., 2020), Little attention has been paid to the spread of the nationwide and even global cities caused by population migration. There is also a literature on how to intervene and control the spread of the COVID-19 virus (Qian et al., 2020; Heymann & Shindo, 2020); Gao et al., 2020).

Although there are many articles on the transmission mechanism of infectious diseases, the research on the law of transmission between regions is still insufficient. Especially with the development of modern transportation and the increasing scale of inter-regional and even transnational population movements, it is becoming increasingly difficult to grasp the spread of infectious diseases across regions.

### 2.2 Research Hypothesis

The transmission factors of the disease are extremely complicated, and it is closely related to the physical conditions of the virus carrier, the environment around, the weather, and the conditions of the contacts. We focus on the influence of spatial connection between cities on the COVID-19 epidemic. The population flow between cities is a key factor affecting the scale of inter-city virus transmission. The population flow is related to urban density and transportation.

Brockmann & Helbing (2013) proposed a new concept of distance, which believes that the spread of disease is not related to the geographical distance between cities, but is closely related to the “effective distance” between cities. What is the effective distance? The effective distance between cities is the length selected by a passenger in various selectable routes from city i to city j. A passenger can be regarded as a random walking particle, who randomly visits the surrounding cities according to the traffic flow. After arriving at the next city, it is converted into a probability to visit the neighboring cities of the next city according to the traffic flow… Finally, the path which the particle most likely to choose from city i to city j is namely the most likely path. The length of this most likely path is the effective distance. We argue that this probabilistic effective distance is related to the flow of population and the degree of transportation convenience, and can reflect the strength of the spatial connection between cities. The effective distance varies with the strength of the spatial connection. This concept is more accurate than traditional connection strength calculations based on the size and distance of the two cities, such as economic connection, market potential, and market access; it is also more accurate than simply using the population flow to characterize the connection, because it cannot express the strength of the connection between the two cities, not to mention the convenience of the connection between the two cities.

With probabilistic effective distances instead of conventional geographic distances, complex spatiotemporal patterns can be simplified into uniform wave transmission patterns. The effective distance is defined as the best path of transportation in the two cities, which depends on the Probabilistic traffic flow.

Assume that *p*_*mn*_ is the conditional probability from node n to destination m is quantified for the migration matrix 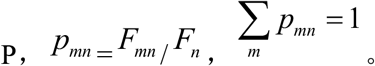。 Weighted links *F*_*mn*_ quantify direct traffic (passengers per day) from node m to node n,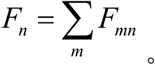 Then,relationship between effective distance and *p*_*mn*_ is:

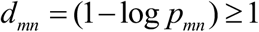

The shorter the effective distance, the greater the probability that the epidemic will spread to the area, then the greater the possibility of increasing the number of imported cases, and the earlier the large-scale epidemic will outbreak (Brockmann & Helbing, 2013). Because healthy individuals are constantly exposed to possible sources of pollution, as long as the lack of protective equipment and effective isolation, they face a serious risk of cross-infection. The reproduction ratio(R0) refers to the average number of secondary infections induced by typical infected individuals in a fully susceptible population. The size of R0 depends on individual contact rate, disease transmission rate, and duration of infection. An increase in R0 can lead to the outbreak of COVID-19 (Read et al., 2020). Based on the classic transmission model and the effective distance of Brockmann & Helbing (2013), the mechanism of epidemic spread and control is shown in Figure 2.

**Figure 1.**
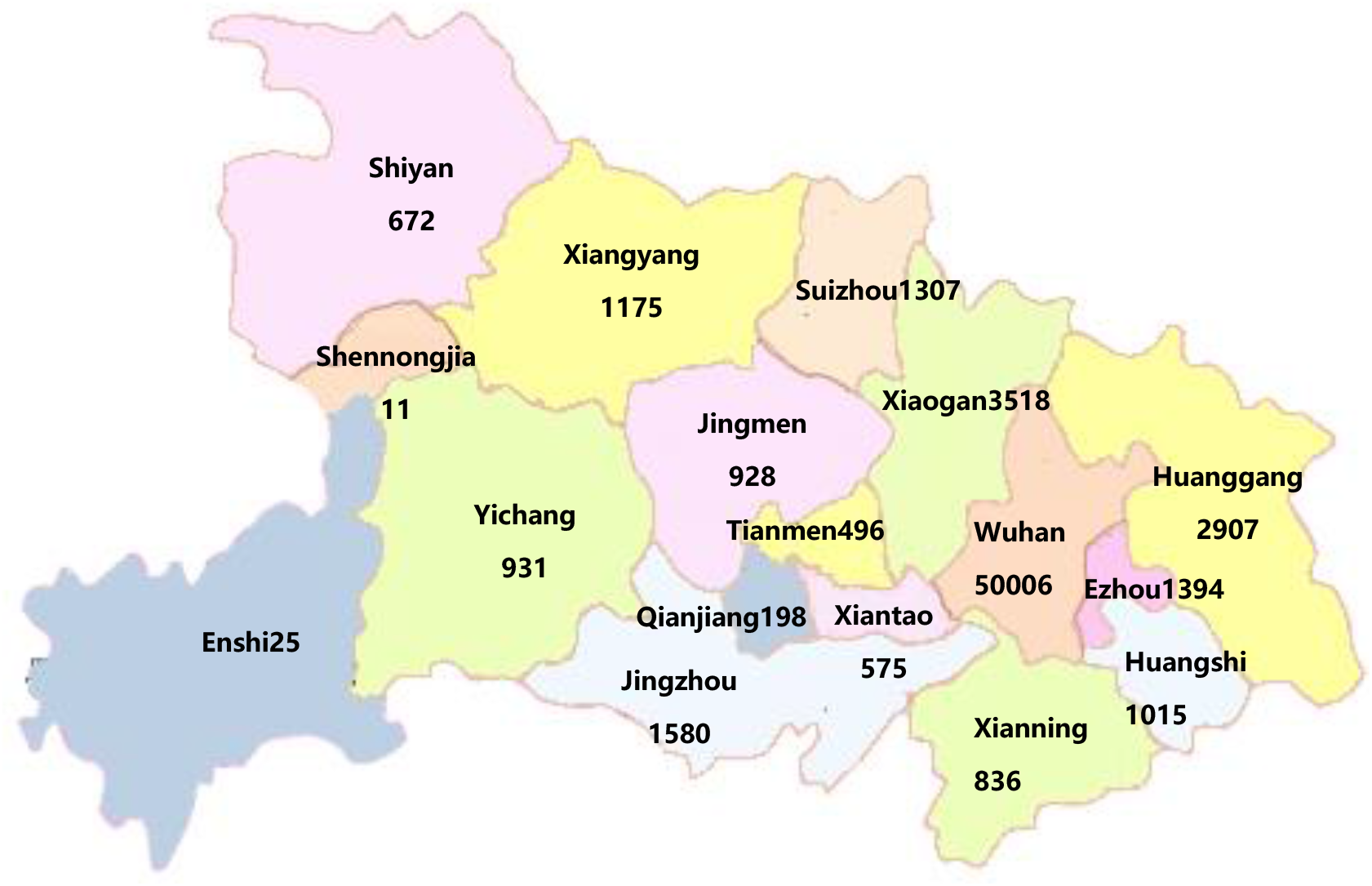
distribution of Cumulative confirmed cases in Hubei province.

**Figure 2.**
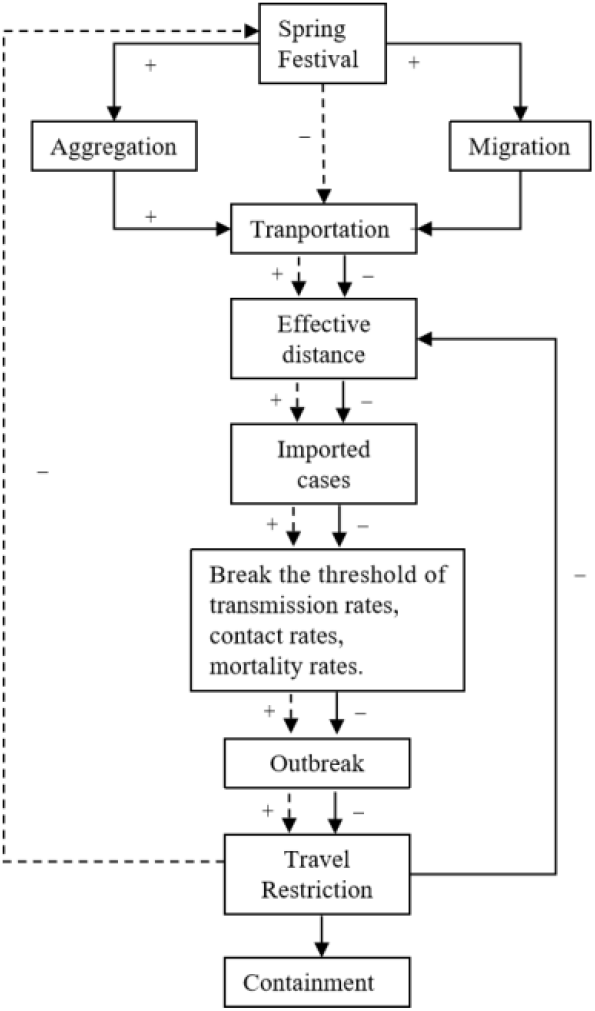
The relationship between effective distance and the transmission of the epidemic.

Based on such understanding of the transmission mechanism, the hypothesis is put forward as follows: the smaller the effective distance, the earlier the outbreak, and the more infectious cases there are in the region.

## 3 methodology

### 3.1 Empirical Model and Variable Descriptions

According to the above mechanism, the empirical model is constructed as follows:

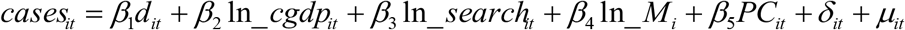

where *cases*_*it*_ is city *i*’ *s* multi-source datasets including cumulative number of confirmed, death, recovered and suspected cases at time t; City *i*’ *s* effective distance *d*_*it*_ at time t as an main explanatory variable, *β*_1_ is the estimated coefficient of *d*_*it*_; if *β*_1_ < 0, effective distance *d*_*it*_ and *cases*_*it*_ coefficient are significant negative correlation; ln_*cgdp*_*it*_, ln_ *search*_*it*_, ln_ *M*_*i*_ and *PC*_*it*_ are control variables at city level, is respectively per capita GDP, Search Index, medical resources(including the number of health institutions, the number of beds per thousand people and the number of health technicians), and prevention and control intensity; *δ*_*it*_ is the time-city fixed effects.

### 3.2 Data Sources

There are five kinds of data sources:

1. Human Migration Big Data^1^. Reflects scale of emigration and immigration population. Under the background of obtaining big data of Human Migration based on geographic location service, we can accurately analyze the destination, specific flow and impact on the epidemic of large-scale Human Migration. Baidu migration data calculates and analyzes the number of intelligent end-users whose positions have changed in 8 hours by comparing the changes of users’ positions, covering all kinds of transportation vehicles such as railway, highway and aviation. The daily numbers of domestic passengers were obtained from the location-based services database of Human Migration Data from Jan.1st, 2020 to Feb.18th, 2020, see http://qianxi.baidu.com/ (in Chinese).
2. Baidu Search Index. Baidu Search Index will reflect a keyword search volume, and with the keyword related to “pneumonia”, “syndromes”, “mask”, “COVID-19” and “travel restriction” will also appear in our search index. The search index can be used to judge the self-consciousness of citizens and explore their behavior trends. Based on the official website of health commission of Hubei Province (wjw.hubei.gov.cn), we obtained the trend of pneumonia epidemic with novel coronavirus infection over time in Hubei Province.
3. The history epidemic date and the trend of pneumonia epidemic with COVID-19 infection are obtained from the official Health Commission. According to regional health policy and measures of disease control and prevention, we evaluate the prevention and control intensity of cities by scoring mechanism. The earlier the policy is issued, the more strict the policy is, the more extensive the monitoring is, and the higher the score is.
4. Statistical Yearbook of Chinese cities for 2019 and Statistical Communique on the 2019 national economic and social development of the city. GDP, population, per capita GDP^1^, the number of health institutions, the number of beds per thousand people and the number of health technicians are obtained from bureau of statistics^2^. If there is no data, the preliminary accounting data released by the provincial (city) bureau of statistics will replace it^3^.

### 3.3 Calculation of Main Variables

#### 3.3.1 Population Migration

Proportion of emigration from other cites: the ratio of emigration from one cites in the overall emigration form all the other cities.

Proportion of immigration to other cites:the ratio of immigration to one cites in the overall emigration to all the other cites.

According to the proportion of emigration from Wuhan since January 1, 2020, it is found that most of the emigration population enters Hubei Province, Hunan Province, Jiangxi Province, Henan Province and Anhui Province. After cutting off the transmission of the virus, the proportion of moving out of Wuhan has decreased significantly. According to the migration big data of Baidu map, the cities with the largest emigration population before New Year’s Eve in 2020 are the Pearl River Delta, Yangtze River Delta and Chengdu-Chongqing urban agglomerations. Shenzhen, Beijing, Shanghai, Guangzhou and Chengdu are the top five cities with the largest emigration population. In addition, Dongguan, Zhengzhou, Hangzhou, Xi’an and other cities are also cities with a large outflow of people before the Spring Festival. Recently, the work resumption was promoted orderly, and the number of people coming to the other places has increased gradually. The top five cities were Shanghai (4.58%), Shenzhen (3.73%), Chengdu (3.64%), Guangzhou (3.56%) and Beijing (3.06%). With the further migration return, central cities such as Shanghai, Shenzhen, Guangzhou, Chengdu, Chongqing and Beijing will face obvious pressure of prevention and control, and measures must be taken in advance to response to the large-scale migration.

#### 3.3.2 Effective Distance

Let *P*_*mn*_ is the fraction of travelers that leave node n and arrive at node m, the effective distance *d*_*nm*_ from a node n to a connected node m is 1 − log*P*_*mn*_, which is generally asymmetric *d*_*mn*_ ≠ *d*_*nm*_.

On the basis of effective distance, we can define the directed length λ(г) of an ordered path г = {*n*_1_,…, *n*_*L*_} as the sum of effective lengths along the path. Moreover, we define the effective distance *D*_*mn*_ from an arbitrary reference node n to another node m in the network by the length of the shortest path from n to m: 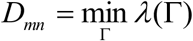 and *D*_*mn*_≠ *D*_*nm*_. From the perspective of a chosen origin node n, the set of shortest paths to all other nodes constitutes a shortest path tree (Brockmann & Helbing, 2013).

Figure 4 shows the effective distance calculated according to the proportion of the population flow from Wuhan to other places. We can see that the cities with the shortest effective distance are all in Hubei province.

**Figure 3.**
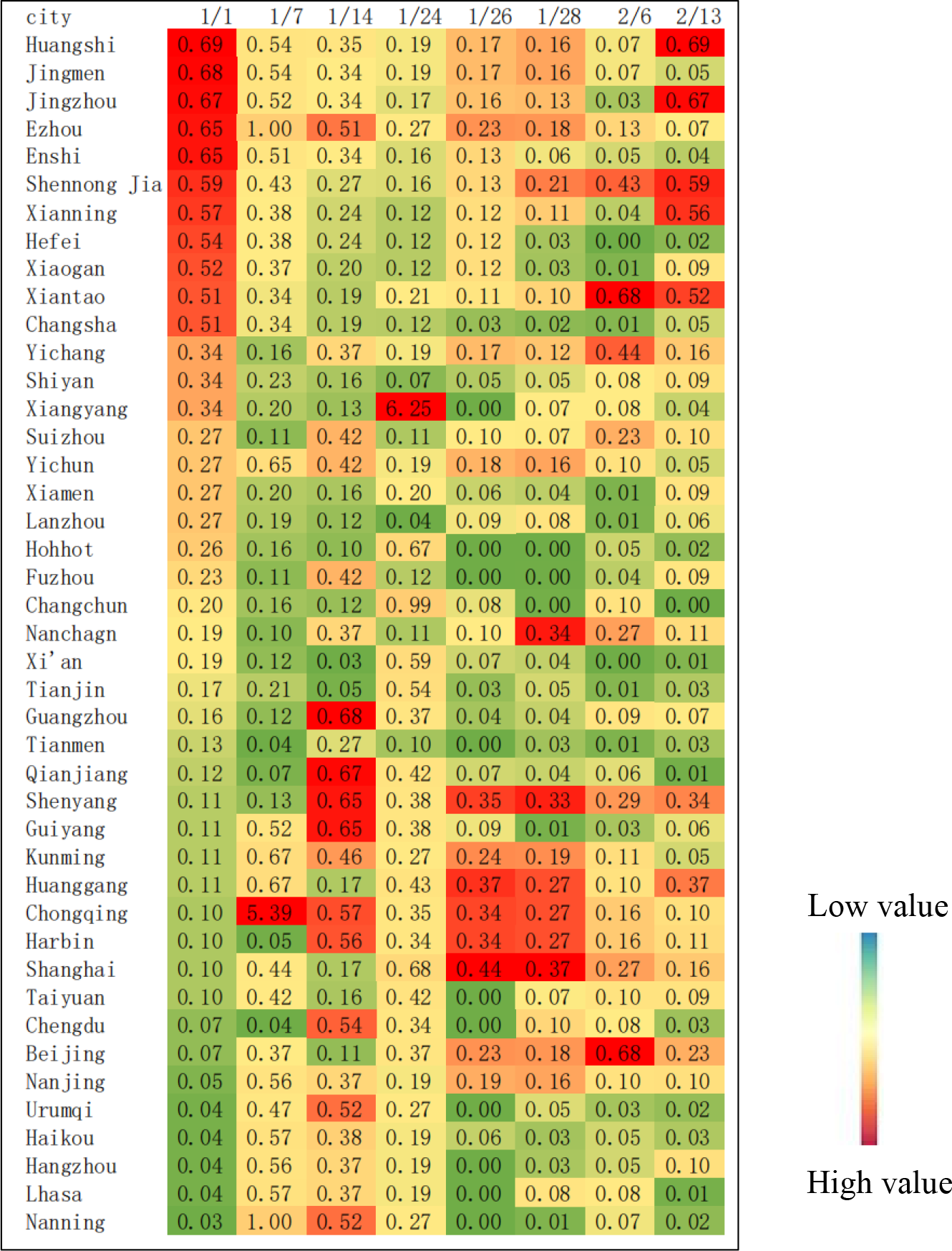
Proportion of immigration to other cites.

**Figure 4.**
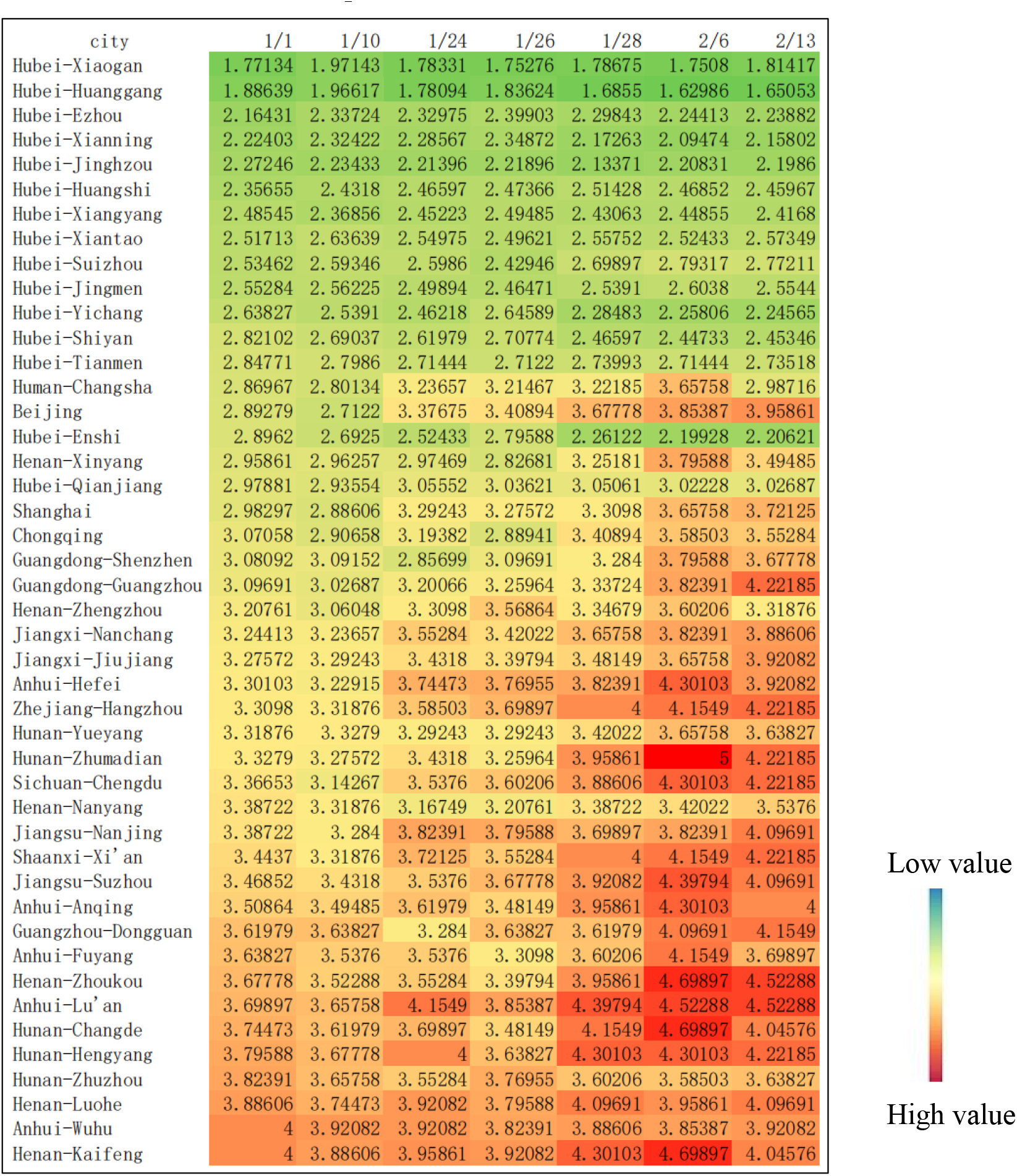
Effective distance from Wuhan to other cities.

#### 3.3.3 Cases of Epidemic

We assess the severity of COVID-19 by utilizing multisource datasets including cumulative and new cases of reported, death, cured and so on. Visualizing the Progression of the COVID-19 Outbreak see figure 6.

**Figure 5.**
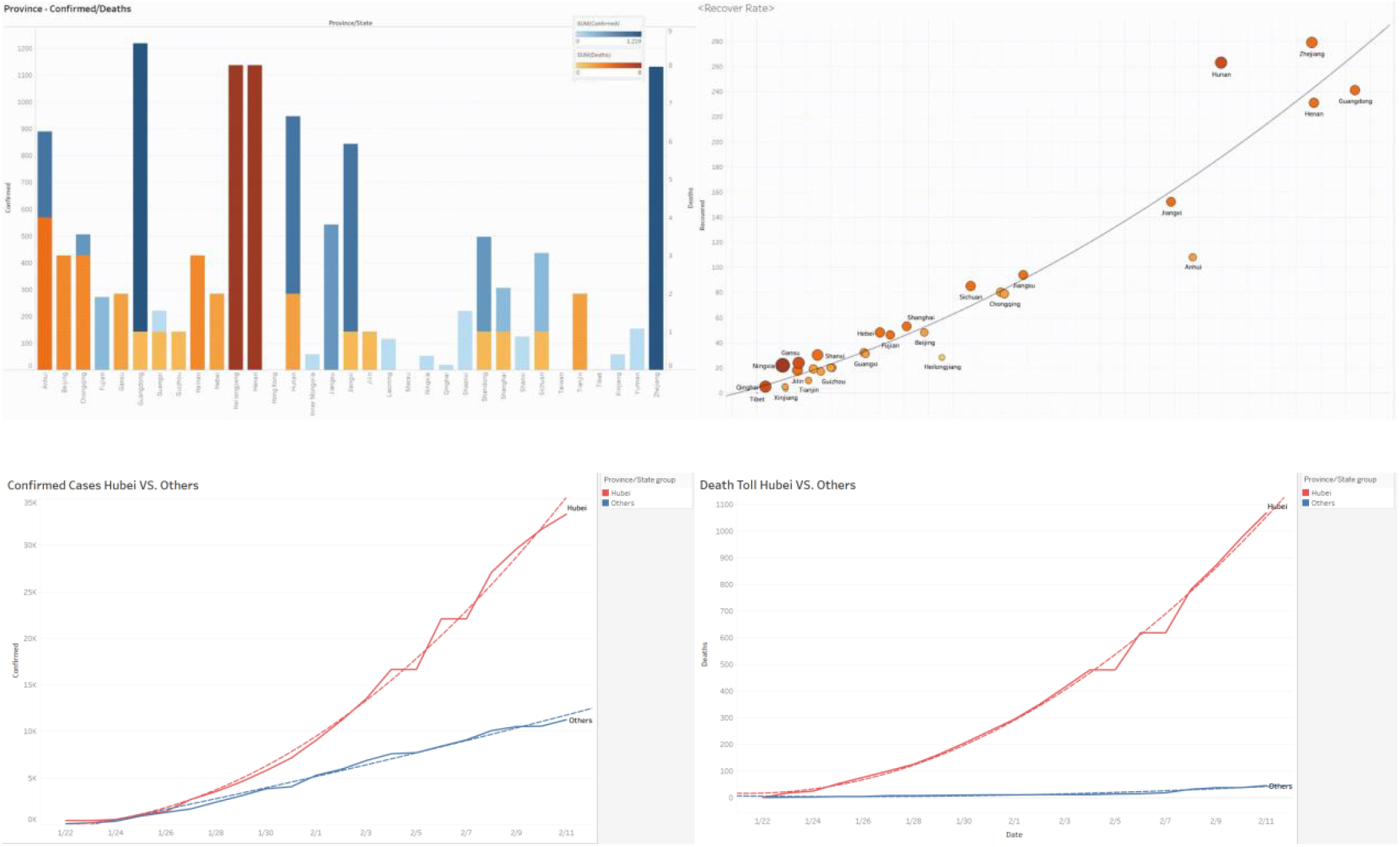
Visualizing the Progression of the COVID-19 Outbreak.

**Figure 6.**
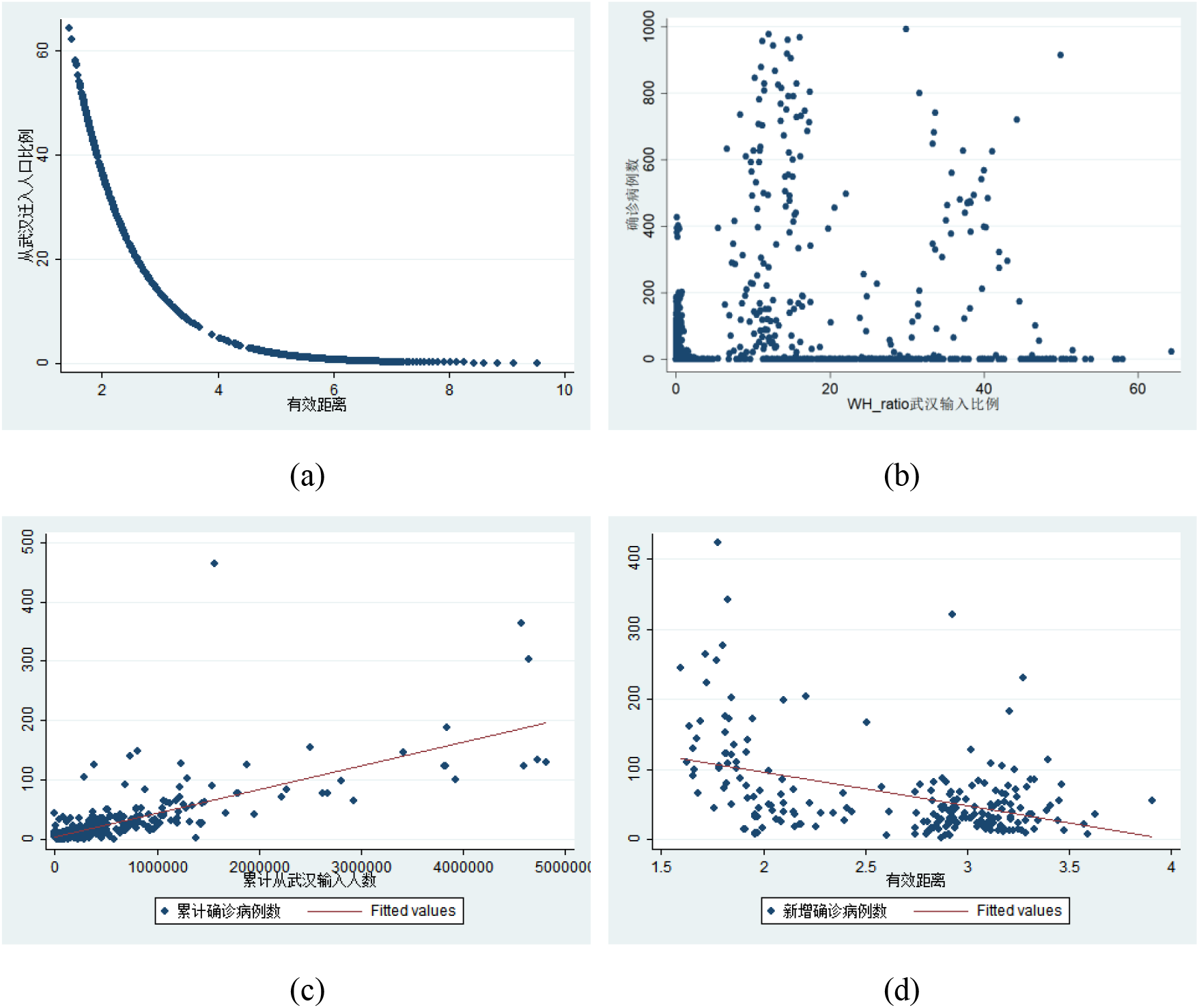
**(a) shows the relationship between the effective distance and emigration; (b) shows the relationship between the cases and emigration in a scatter plot without considering the incubation period; (c) and (d) shows the fitting results considering the impact factor of the incubation period**.

Confirmed rate per million people: the number of confirmed cases for every million people.

Cure rate for confirmed cases:how many people of the confirmed cases were cured and discharged,reflects the quality of medical treatment and cure condition.

Mortality rate for confirmed cases: how many people of the confirmed cases have died, reflects the severity of the epidemic in this area.

Diffusion index: the number of newly infirmed patients on that day / the cumulative number of infirmed on the previous day.

Reduction index: (currently new cures + new deaths on the day) / cumulative confirmed cases on the previous day.

#### 3.3.4 Prevention and Control Intensity

We establish an evaluation system for the intensity of epidemic prevention and control, including: launching level 1 response, stop of inter-city passenger transportation, stop of ordinary public transportation in the city, stop of part of public places in the city, closed management of residential areas, centralized isolation of returnees from the key epidemic areas (Hubei) for 14 days, centralized isolation of returnees from non-key epidemic areas for 14 days, prohibition of local personnel to go out, centralized isolation of close contact of confirmed patients for 14 days, isolation of suspected patients for medical observation and treatment. Each item is divided into full, partial, and no three items, with a score of 1, 0.5, and 0 respectively. The content and implementation time of the measures come from the information or announcements issued by epidemic prevention and control headquarters in prefectural-level districts.

### 3.4 statistical description

We will explore the relationship between effective distance from Wuhan and the number of COVID-19 cases confirmed in different cities. As shown in Figure 6(b), the correlation between the proportion of the outflow of people from Wuhan and the number of cases is not significant on the whole. We analyze that there are two reasons cause the result: First, he incubation period of COVID-19 is long, the patients only have mild symptoms, such as fever, fatigue and cough. The paper titled “Time-varying transmission dynamics of Novel Coronavirus Pneumonia in China “published in the international medical authoritative journal “the New England Journal of Medicine” revealed that the average incubation period of new coronary pneumonia is 5.2 days (Li et al., 2020); Second is that the first cases reported in most cities are after January 20. Although the number of cases was not previously reported, the possibility of an earlier infection cannot be ruled out. It is estimated that the outbreak started much earlier, and both within China and international infectious exports occurred before January and in early January. Based on the analysis above, this paper considers the incubation period, and replaces the actual time of infection with the time of cases confirmed, and obtains the relationship between the flow of people out of the Wuhan and the number of pneumonia cases in the other cities, as shown in Figure 6.

From the data and figures above, we can see that if we take the incubation period into consideration, the cumulative confirmed cases are significantly related to the outflow of people from Wuhan, and the effect of effective distance on the new confirmed cases is significantly negative.

## 4 Empirical Analysis

### 4.1 Benchmark Regression

According to the selection of the model and calculation the variable, the benchmark regression results are shown in Table 2.

**Table 1.** Regional distribution of COVID - 19 cases in mainland China as of 20 March.

|  | Cumulative<br>Confirmed<br>Case | Proportion in<br>Mainland China<br>(%) | Confirmed<br>Deaths | Proportion in<br>Mainland China<br>(%) | Existing<br>Confirmed<br>Cases | Proportion in<br>Mainland China<br>(%) |
| --- | --- | --- | --- | --- | --- | --- |
| Hubei | 67801 | 83.4 | 3139 | 95.5 | 3431 | 86.9 |
| among:<br>Wuhan | 50006 | 61.5 | 2531 | 77.0 | 3407 | 86.3 |
| Areas<br>outside<br>Hubei | 13484 | 16.6 | 148 | 4.5 | 516 | 13.1 |
| Total | 81285 | 100.0 | 3287 | 100.0 | 3947 | 100.0 |

Source: website of National Health Commission

**Table 2.** statistical description.

| Variable | Obs | Mean | Std. Dev. | Min | Max |
| --- | --- | --- | --- | --- | --- |
| Cumulative Cases | 14314 | 28.976 | 222.838 | 0 | 13910 |
| New cases | 14008 | 1.949 | 15.670 | 0 | 1252 |
| Deaths cases | 14216 | 0.325 | 3.121 | 0 | 85 |
| Cure cases | 14240 | 3.758 | 24.186 | 0 | 1059 |
| GDP | 14328 | 2963.587 | 40022.274 | 164 | 38155.3 |
| Population | 14328 | 434.267 | 292.914 | 55.44 | 2416 |
| Per capita GDP | 14328 | 6.021 | 3.629 | 1.077 | 20.671 |
| Medical institutions | 14328 | 2642.938 | 2074.349 | 330 | 11196 |
| Search Index | 14229 | 137.060 | 168.177 | 0 | 2627 |
| Prevention and control intensity | 14328 | 3.592 | 3.879 | 0 | 10 |
| Emigration | 14328 | 8.421 | 38.627 | 0 | 84.834 |
| Effective distance | 14328 | 6.709 | 1.421 | 1.440 | 9.517 |

**Table 2.** benchmark regression.

|  | (1) | (2) | (3) | (4) | (5) |
| --- | --- | --- | --- | --- | --- |
| Variables | Cumulative cases | Cumulative cases per thousand | Death cases | Death cases per thousand | New cases per thousand |
| Effective distance | -56.843**<br>(-2.33) | -1.65***<br>(-2.90) | -1.335**<br>(-2.34) | -0.040**<br>(-2.26) | -0.219***<br>(-3.15) |
| Per capita GDP | -314.602***<br>(-2.92) | -9.838***<br>(-2.96) | -6.623**<br>(-2.42) | -0.200**<br>(-2.28) | -1.040***<br>(-3.33) |
| Search Index | -88.837**<br>(-2.69) | -2.032***<br>(-3.40) | -0.841***<br>(-2.79) | -0.029**<br>(-2.36) | -0.183***<br>(-3.48) |
| The number of health institutions | -107.419**<br>(-2.30) | -3.417**<br>(-2.53) | -3.122**<br>(-2.37) | -0.091**<br>(-2.27) | -0.501***<br>(-3.18) |
| Prevention and control intensity | -25.199**<br>(-2.01) | -0.644**<br>(-2.52) | -0.488**<br>(-2.10) | -0.011***<br>(-2.62) | -0.036***<br>(-2.59) |
| Time fixed effect | Yes | Yes | Yes | Yes | Yes |
| City fixed effect | Yes | Yes | Yes | Yes | Yes |
| Observation | 3466 | 3466 | 4233 | 4233 | 4254 |
| R2 | 0.608 | 0.604 | 0.487 | 0.490 | 0.494 |

\*\*\*p<0.01, \*\* p<0.05, \* p<0.1

Table 2 reports the benchmark regression results of the effect of effective distance on the number of new cases, in which control variables, time and city fixed effect is added, and the regression coefficient of effective distance is negative significantly, which shows that when other conditions are constant, the shorter the effective distance, the more the cumulative number of confirmed cases, the number of confirmed cases per thousand, cases of deaths, the cases deaths per thousand, and the number of newly confirmed cases per thousand there are. It is worth noting that the greater the intensity of urban epidemic prevention and control, the fewer new confirmed cases, indicating the effectiveness and necessity of various prevention and control measures. The more health institutions there are, the fewer cases there are. In the model, some urban characteristic factors, including per capita GDP, beds and health technicians per thousand have a negative impact on the number of new cases, indicating the rationality of the model. In the model, per capita GDP has a negative impact on the number of new cases, indicating that the higher the GDP in a region is, the more the population flow is, and the higher the number of new cases is. The search index has a significant negative impact on the number of outbreaks, indicating that the more severe the outbreak, the higher the awareness of how to protect.

### 4.2 Robustness

In this paper, a variety of methods are used to test the robustness of the benchmark regression conclusion, shown in Table 3.

**Table 3.** The results of Robust test.

|  | (1) Proxy variables<br>for explanatory<br>variable | (2) PPML | (3) Proxy variables<br>for explained<br>variable | (4) Temporal Clustering |  |
| --- | --- | --- | --- | --- | --- |
| Variables | New cases | New cases | New cases | New cases | New cases |
| Emigration |  |  | 2.471* (1.70) |  |  |
| Effective distance | -5.998** (-2.51) | -0.339***<br>(-4.15) |  | -9.682***<br>(-2.68) | -1.275**<br>(-2.39) |
| Per capita GDP | -24.618*** (-2.87) | -5.430***<br>(-3.24) | -7.166<br>(-1.37) | -4.397<br>(-0.25) | -6.405**<br>(-2.03) |
| Search Index | -8.003** (-2.29) | -0.769***<br>(-3.65) | -9.027**<br>(-2.23) | -11.013***<br>(-2.61) | -1.440*<br>(1.82) |
| The number of<br>health institutions | -3.956***<br>(-3.01) | -0.681***<br>(-2.99) | -4.079<br>(-0.97) | 25.299***<br>(2.89) | 14.205***<br>(-2.93) |
| Prevention and | -0.925* (-1.76) | -0156*** | -1.045* | -0.854 | -34.707* |
| control intensity |  | (-2.79) | (0.069) | (-0.51) | (-1.85) |
| Time fixed effect | YES | YES | YES | YES | YES |
| Entity fixed effect | YES | YES | YES | YES | YES |
| Observation | 3406 | 3406 | 3406 | 395 | 1349 |
| R2 | 0.570 | 0.580 | 0.560 | 0.773 | 0.755 |

\*\*\*p<0.01, \*\* p<0.05, \* p<0.1

1. Using multi-source datasets including cumulative number of reported, death, quarantined and suspected cases, cure rate for confirmed cases, mortality rate for confirmed cases as the explanatory variable, the result is also robust.
2. VPPML method. Silva & tennero (2006) pointed out that if there is an option of heteroscedasticity or zero value (due to the fact that the number of initial case in many cities is zero), Poisson maximum likelihood model (PPML) should be used to test the robustness.
3. The effective distance is replaced by the absolute number of people moving out of Wuhan. the explained variable. After replacing explanatory variable, the regression result is also robust.
4. (4)Different Sample time selection. The sample period of standard regression is from January 1st, 2020 to February 18th, 2020. However, there are still many events that may have a significant impact on the number of cases during the sample period, such as the Spring Festival and lockdown of the city. In order to test whether the choice of time period has an impact on the regression results, further regression is carried out according to the sub-samples before and after Wuhan has been locked down. In each sub-sample, the effect of effective distance on the number of new confirmed cases is always negative, which implies that results are robust.

### 4.3 Endogeneity Test

The incubation period of the novel coronavirus is the main source of endogeneity. In this paper, we use the lag of effective distance as an instrument variable to address the endogeneity, which is shown in table 4.

**Table 4.**
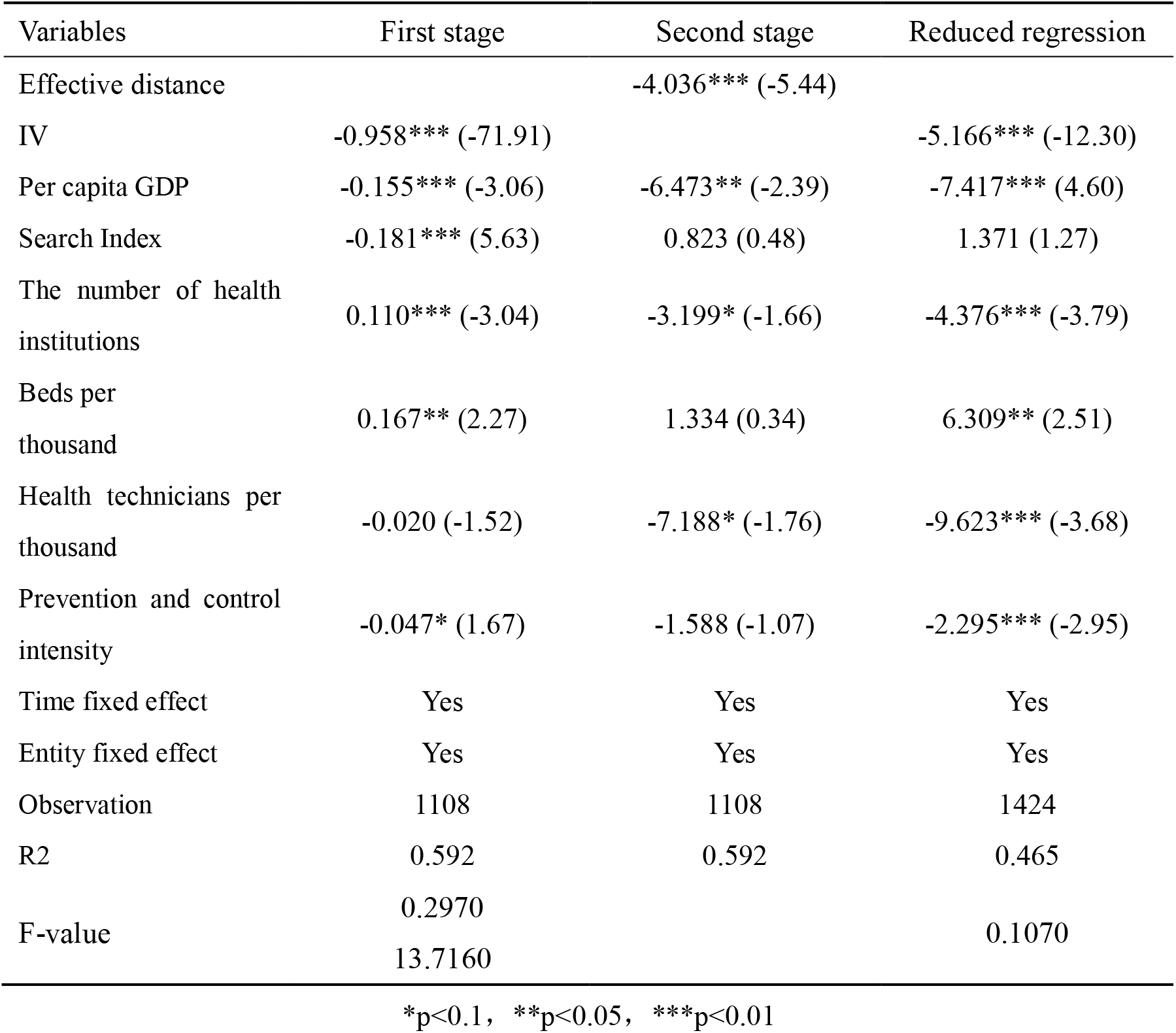
Instrument variables regression result.

## 5 Conclusions and implications

In this article, by introducing the effective distance to reflect the strength of the connection between cities and constructing an empirical model. it is concluded that: (1) the scale of local COVID-19 epidemic is closely related to the effective distance, that is, the strength of the connection between the two cities. The shorter the effective distance, the more newly confirmed cases and the more severe the local epidemic; (2) The greater the prevention and control efforts of urban epidemic, the more health institutions, the fewer cases there are; (3) The regions with higher per capita GDP have more population flow and more cases of epidemic cases; The more severe the outbreak, the higher the awareness of how to protect.

According to the conclusions above, there are the following implications:

First, strengthen cross-regional epidemic prevention and control linkages to achieve joint prevention and control between regions. The smaller the effective distance, the more convenient the connection between the two cities, the larger the scale of population flow, so the easier the epidemic will spread across regions, the more important it is for cross-regional cooperation to prevent and control the epidemic. In the future epidemic prevention and control, it’s necessary to establish cross-region epidemic information sharing, prevention and control measures linkage, and medical resources collaboration.

Second, strengthen comprehensive measures for comprehensive prevention and control of the epidemic.

The large investment in prevention and control which has a large impact on the economy and society can control the epidemic in a short time and shorten the time of the impact of the epidemic on the economy and society. This requires comprehensive measures. On the one hand, the government, society, and residents should make concerted efforts to jointly control, accelerating the speed of epidemic control; on the other hand, we must strengthen the multi-faceted reserves of materials needed for epidemic prevention and control.

the government, society, and residents jointly control and control, and work together to accelerate the speed of epidemic control; On the other hand, it is necessary to strengthen the multi-faceted stock of materials needed for epidemic prevention and control, so that comprehensive measures can be taken when the epidemic occurs

Third, investment in public health resources should be strengthened. The more developed the city is, the easier it is for people to contact, and the larger the scale of population flow, the easier it is for the epidemic to spread. Therefore, the more developed the economy, the more it is necessary to increase investment in public health resources.

## Data Availability

all data included in this study are available upon request by contact with the corresponding author.

## Footnotes

1 The urban administrative division is adopted for the urban migration boundary, including the districts, counties, townships and villages under the jurisdiction of the city.

1 GDP, added value of various industries and per capita GDP are calculated at current prices, and the growth rate is calculated at comparable prices.

2 The average exchange rate of US dollar in 2019 is about 6.89;

3 The GDP of Beihai is the reciprocal deduction = the initial check number of Guangxi Region - the initial check number or estimated number of other cities. The data in some bulletins are preliminary statistics. Due to the rounding, some data are unequal in total and sub total.

